# HIV Risk and Intention to use PrEP among Sexually Active Female University Students in Zambia: A Cross-Sectional Survey to Understand Influential Factors

**DOI:** 10.1101/2024.12.12.24318948

**Authors:** K Hampanda, M Bolt, L Nayame, T Hamoonga, M Sehrt, J Thorne, M Harrison, J Pintye, A Amstutz, L Abuogi, O Mweemba

## Abstract

**Background:** Limited research exists on pre-exposure prophylaxis (PrEP) interest or use among female university students in high HIV-prevalence African settings. This study sought to establish the relationship between epidemiologic and perceived HIV risk and PrEP intention among Zambian female university students.

**Methods:** We recruited female students at an urban university to complete a survey on intention to use PrEP in the next year (primary outcome); other PrEP knowledge, attitudes, and behaviors; demographics; epidemiologic HIV risk and risk perception. Descriptive statistics, regression and mediation analyses were used.

**Results:** Of the 454 sexually active participants, 118 (26%) reported PrEP intention. Actual PrEP use was rare (< 5%). The odds of PrEP intention increased for those with perceived high HIV risk (aOR 3.08; 95% CI 1.71-5.55) and with each year at university (aOR 1.47; 95% CI 1.21-1.80) but decreased with higher PrEP stigma (aOR 0.91; 95% CI 0.86-0.96) and more negative PrEP perceptions (aOR 0.91; 95% CI 0.85-0.97). More epidemiologic risk factors were originally associated with PrEP intention (OR 1.24; 95% CI 1.01-1.53 for each risk factor), though this relationship weakened after adjustment for perceived HIV risk, which mediated 69% of the relationship between epidemiologic HIV risk and PrEP intention. Only 29% of high-risk participants recognized their high epidemiological HIV risk (3+ risk factors).

**Conclusions:** Along with PrEP education and stigma reduction, there is a need for approaches that help female university students in Zambia accurately identify their HIV risk to make informed PrEP decisions.

## INTRODUCTION

In Zambia, HIV prevalence is four times higher among young women (9%) compared to young men (2%) ages 20 to 24 years. ^1^ Female university students in high HIV prevalence settings of Southern Africa, such as Zambia, are a neglected, high-risk sub-population for HIV acquisition.^2-4^ Out-of-school adolescent girls and young women (AGYW) have often been the target of oral pre-exposure prophylaxis (PrEP) promotion.^5,6^ There is growing recognition that in-school AGYW have similar risk factors as out-of-school AGYW, and may in fact, have increased risk of HIV exposure^7^ In-school AGYW may have a greater number of lifetime sexual partners due to delayed marriage, and use transactional sex, often with older male partners, to subsidize education costs and living expenses ^8,9^. In Zambia, HIV prevalence is higher among women with higher education (16%) compared to those with no formal education (11%).^7^

Significant investment has been made to increase public education and the number of sites offering oral PrEP in Zambia, as well as create demand among priority populations, including AGYW. The little research that exists with Zambian AGYW indicates relatively high uptake but low continuation of oral PrEP. ^10,11^ Outside of Zambia, programs delivering oral PrEP to AGYW, mainly in South Africa and Kenya, have reported low uptake, and poor adherence and persistence. ^12-17^ There is a dearth of research on PrEP use or intention among female university students in high HIV prevalence settings of Africa, despite high HIV risk. ^18,19^

‘Market segmentation’ of AGYW at high risk of HIV is needed to characterize potential avenues for tailored interventions to improve PrEP uptake and persistence. ^20^ Numerous barriers to PrEP use among African AGYW have been established, including low risk perception, stigma, lack of partner support, and lack of access/delivery options. ^21,22^ Yet, female university students have generally not been included or the focus of this prior research. To fill this gap and plan for a tailored intervention, we conducted a cross-sectional survey at a large university in Lusaka, Zambia. The primary objective of this study was to establish a profile of sexually active female university students who intend to use PrEP. In addition, we examined the relationship between HIV risk perception, epidemiological HIV risk, and PrEP intention to further characterize this understudied sub-population of AGYW.

## METHODS

### Setting

Zambia has a generalized HIV epidemic with a high HIV prevalence (9.9%) among those 15-49 years and annual incidence of 0.34%. ^23^ Lusaka province, the study setting, has the highest HIV prevalence (14.4%) in Zambia^23^ Oral PrEP is widely available in Lusaka free of charge at government-run health centers. The study was approved by relevant ethics committees in the United States and Zambia.

### Sampling and Data Collection

From February to April 2022, we recruited a convenience sample of female university students (n=806). Currently enrolled female-identifying students ≥18 years of age completed an online survey in English (official Zambian language). We partnered with the university’s medical services and peer educators to distribute flyers with the survey QR code/website link. A virtual flyer was distributed to student WhatsApp groups by university administration. After consent and screening questions (age, gender, current student), participants completed the online survey (∼15 min). Data was collected and stored using REDCap. ^24,25^

### Outcome of interest

The primary outcome (intention to use PrEP) was measured by asking “*How likely are you to use PrEP in the next year*?” with a 5-point Likert scale (definitely will not use; probably will not use; unsure; likely will use; definitely will use). The question was dichotomized to reflect those who intended to use PrEP (i.e. “likely/definitely will use”) versus those who did not or were unsure.

### Exposures of interest

Epidemiologic HIV risk was measured using the validated HIV Risk Index, ^26,27^ which indicates AGYW with ≥3 risk factors are 15 times as likely to acquire HIV as those with <3 factors and meet a high-risk HIV threshold. ^27,28^ The index includes sociodemographic characteristics (age 20 to 24 years), sexual behavior and relationship characteristics (2 or more partners in the past year; ever exchanging sex for money or gifts; having recent sexual partner(s) who are 5 years or older; and suspected/known partner concurrency), sexually transmitted infection (STI) symptoms in the past 6 months (abnormal vaginal discharge or genital sores/ulcers), and prior pregnancy. Higher numbers indicate higher HIV risk. We dichotomized these risk scores into high epidemiologic HIV risk (3 or more risk factors) or low HIV risk (0-2 risk factors). Self-perceived HIV risk was measured by asking “*What do you think your risk is of getting HIV in the next year?”*. “No risk at all”, and “small risk” constituted low risk perception, with responses of “50/50 risk”, “high risk” and “very high risk” were labeled high risk perception. Negative PrEP perceptions were measured using established scales with Likert responses. ^29,30^

### Data cleaning and multiple imputation for missing data

Survey deployment of demographics and PrEP intention commenced on February 15, 2022. From March 2, 2022 onward, additional questions on sexual behavior and history were added after an error in the survey skip pattern was identified. Since a substantial proportion of responses for sexual behavior and history were missing, multiple imputation was used to avoid substantial losses of power and biased estimation, as it has shown to be beneficial over complete-case analysis even in situations where the proportion of missing data for a covariate is large. ^31-33^ Multiple imputation was conducted with the mice package in R, using predictive mean matching.^34,35^ Epidemiological risk was calculated after imputation. Year in school was mean-centered for interpretability in regression modeling. Fifty imputations were utilized. A complete-case analysis was conducted for sensitivity analysis (see Appendix). We excluded participants who reported living with HIV, never had sex, or missing PrEP intention data (the primary outcome) from analysis.

### Data analysis

Descriptive statistics (counts and proportions for categorical variables; means and standard deviations for continuous and Likert scale variables) were calculated for the original, pre-imputed data (i.e. complete case). All subsequent analyses were performed with multiply imputed datasets. Logistic regression was used for all regression modeling. Significant (alpha = .05) variables in unadjusted models were included in multivariable models. Regression results from all imputed datasets were pooled together to create final parameter estimates using Rubin’s rules. ^36^ All data cleaning and analyses were conducted in R (version 4.4.0) and associated R packages. ^34,37-43^

To evaluate the presence and magnitude of the relationship between epidemiological risk and intention of using PrEP mediated by perceived HIV risk, we followed classic mediation framework proposed by Baron and Kenny (1986). We fitted models between the predictor and mediator, and between the outcome with the predictor and mediator together, then following the recommendations of Rijnhart et al. (2019) to a*b to calculate the indirect effect and a*b/(ab + c’) to calculate the proportion mediated (all letters represent log-odds of the appropriate mediation model pathways). ^44,45^

To evaluate whether participants accurately assessed their true HIV risk, McNemar’s tests were performed on all imputed datasets on cross tabulations of dichotomized perceived HIV risk and dichotomized epidemiological risk.

## RESULTS

Of 806 total survey responses, 454 sexually active (i.e., HIV at-risk) female students not living with HIV met final inclusion criteria. Table 1 presents the sample characteristics. The average age was 22.6 years old (SD 2.6) with 10% financially comfortable. Most (98%) were born in Zambia though 56% were born outside of Lusaka Province. Most (78%) were in their first to third year of university. Most (52%) lived on campus or in a rental house/room (36%). Very few were currently taking PrEP (<1%) or ever used PrEP (4.7%). Few (22%) knew someone taking PrEP. The majority (81%) perceived they were low HIV. Most (70%) reported knowing their last sexual partner’s HIV status.

**Table 1:** Sample characteristics.

|  |  | Likelihood of using PrEP |  |
| --- | --- | --- | --- |
| Characteristic | Overall, N = 454<sup>1</sup> | Likely, N = 118<sup>1</sup> | Unlikely, N = 336<sup>1</sup> |
| General demographics |  |  |  |
| Age | 22.57 (2.56) | 22.55 (3.27) | 22.58 (2.27) |
| (Missing) | 2 | 1 | 1 |
| Financial situation<sup>2</sup> |  |  |  |
| Comfortable | 43 (10.0%) | 15 (14%) | 28 (8.7%) |
| Poor | 389 (90%) | 95 (86%) | 294 (91%) |
| (Missing) | 22 | 8 | 14 |
| Location of birth |  |  |  |
| Lusaka province | 192 (42%) | 48 (41%) | 144 (43%) |
| Zambia | 255 (56%) | 70 (59%) | 185 (55%) |
| International | 5 (1.1%) | 0 (0%) | 5 (1.5%) |
| (Missing) | 2 | 0 | 2 |
| Year at University |  |  |  |
| 1st year | 123 (27%) | 45 (39%) | 78 (23%) |
| 2nd year | 101 (22%) | 28 (24%) | 73 (22%) |
| 3rd year | 129 (29%) | 33 (28%) | 96 (29%) |
| 4th year | 65 (14%) | 9 (7.8%) | 56 (17%) |
| 5th year | 22 (4.9%) | 0 (0%) | 22 (6.6%) |
| 6th year or more | 11 (2.4%) | 1 (0.9%) | 10 (3.0%) |
| (Missing) | 3 | 2 | 1 |
| Living space |  |  |  |
| A rental house/room | 143 (36%) | 47 (46%) | 96 (33%) |
| Free housing with friend/relative | 36 (9.1%) | 10 (9.8%) | 26 (8.9%) |
| On campus | 204 (52%) | 42 (41%) | 162 (55%) |
| Own home | 12 (3.0%) | 3 (2.9%) | 9 (3.1%) |
| (Missing) | 59 | 16 | 43 |
| Familiarity with PrEP |  |  |  |
| Currently taking PrEP | 4 (0.9%) | 3 (2.6%) | 1 (0.3%) |
| (Missing) | 6 | 3 | 3 |
| Has ever take PrEP before | 21 (4.7%) | 12 (10%) | 9 (2.7%) |
| (Missing) | 9 | 2 | 7 |
| Know someone taking PrEP | 97 (22%) | 36 (32%) | 61 (19%) |
| (Missing) | 22 | 7 | 15 |
| HIV and birth control familiarity |  |  |  |
| Perceived HIV risk |  |  |  |
| Low risk | 324 (81%) | 65 (63%) | 259 (88%) |
| High risk | 76 (19%) | 39 (38%) | 37 (13%) |
| (Missing) | 54 | 14 | 40 |
| Thought about getting HIV | 237 (54%) | 67 (58%) | 170 (52%) |
| (Missing) | 14 | 2 | 12 |
| Knows HIV status of last partner |  |  |  |
| Knew partner's HIV status | 316 (70%) | 76 (64%) | 240 (71%) |
| Did not know | 138 (30%) | 42 (36%) | 96 (29%) |
| Birth control used |  |  |  |
| Condoms | 317 (73%) | 84 (75%) | 233 (73%) |
| Other modern method | 69 (16%) | 21 (19%) | 48 (15%) |
| Traditional method | 17 (3.9%) | 2 (1.8%) | 15 (4.7%) |
| Don't use birth control | 29 (6.7%) | 5 (4.5%) | 24 (7.5%) |
| (Missing) | 22 | 6 | 16 |
| **Self-identified epidemiological risk factors** |  |  |  |
| Age 20 - 24 | 347 (76%) | 87 (74%) | 260 (77%) |
| Number of sex partners in last year | 1.59 (1.79) | 1.76 (1.49) | 1.53 (1.88) |
| (Missing) | 236 | 64 | 172 |
| Had sex for money | 17 (8.0%) | 9 (17%) | 8 (5.1%) |
| (Missing) | 242 | 64 | 178 |
| Had a partner 5 years older | 82 (38%) | 24 (45%) | 58 (36%) |
| (Missing) | 241 | 65 | 176 |
| Know/suspect partner has other partners within 6 mo | 153 (50%) | 52 (61%) | 101 (45%) |
| (Missing) | 146 | 33 | 113 |
| STI symptoms in last 6 months | 74 (17%) | 26 (23%) | 48 (15%) |
| (Missing) | 9 | 3 | 6 |
| Have been pregnant | 78 (18%) | 19 (17%) | 59 (18%) |
| (Missing) | 9 | 3 | 6 |
| Number of risk factors | 2.33 (1.44) | 2.61 (1.66) | 2.22 (1.35) |
| (Missing) | 320 | 82 | 238 |
| Dichotomous risk variable |  |  |  |
| Low risk | 83 (62%) | 21 (58%) | 62 (63%) |
| High risk | 51 (38%) | 15 (42%) | 36 (37%) |
| (Missing) | 320 | 82 | 238 |
| Any risk factors |  |  |  |
| No risk factors | 7 (5.2%) | 1 (2.8%) | 6 (6.1%) |
| At least 1 risk factor | 127 (95%) | 35 (97%) | 92 (94%) |
| (Missing) | 320 | 82 | 238 |

1 Mean (SD); n (%)
2 Question asked: 'How would you classify your financial situation these days?'

On average, participants without missing data reported 2.33 epidemiologic HIV risk factors (SD 1.44) and 1.6 sexual partners in the past year. Thirty eight percent reported having a sexual partner 5 years or older. Half knew or believed that their sexual partner had other sexual partners. Seventeen percent reported having STI symptoms in the past 6 months and 18% reported ever being pregnant. Almost all (95%) had at least one risk factor for HIV while 38% had ≥3 risk factors (i.e., high epidemiologic HIV risk), though the high proportion of the sample having at least one risk factor is partly explained by 76% being 20-24 years of age ^26^.

Table 2 presents endorsement of each item and overall scores for the stigma and negative PrEP perceptions scales. With scales ranging from 0 (completely disagree) to 4 (completely agree), on average, there were low levels of negative PrEP perceptions (item scores of 0.49 - 1.99) and stigma (item scores of 0.83 - 2.07).

**Table 2:**
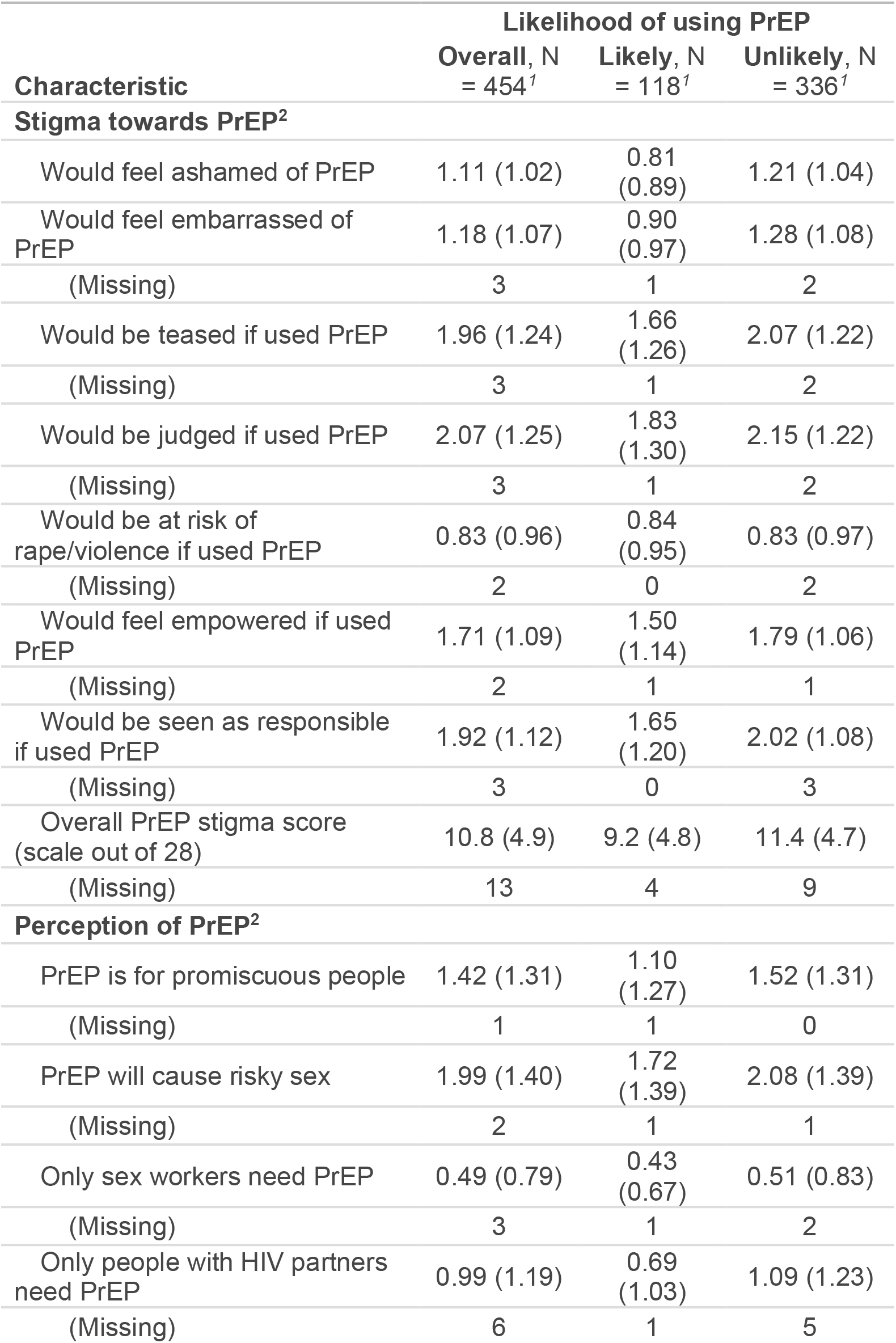
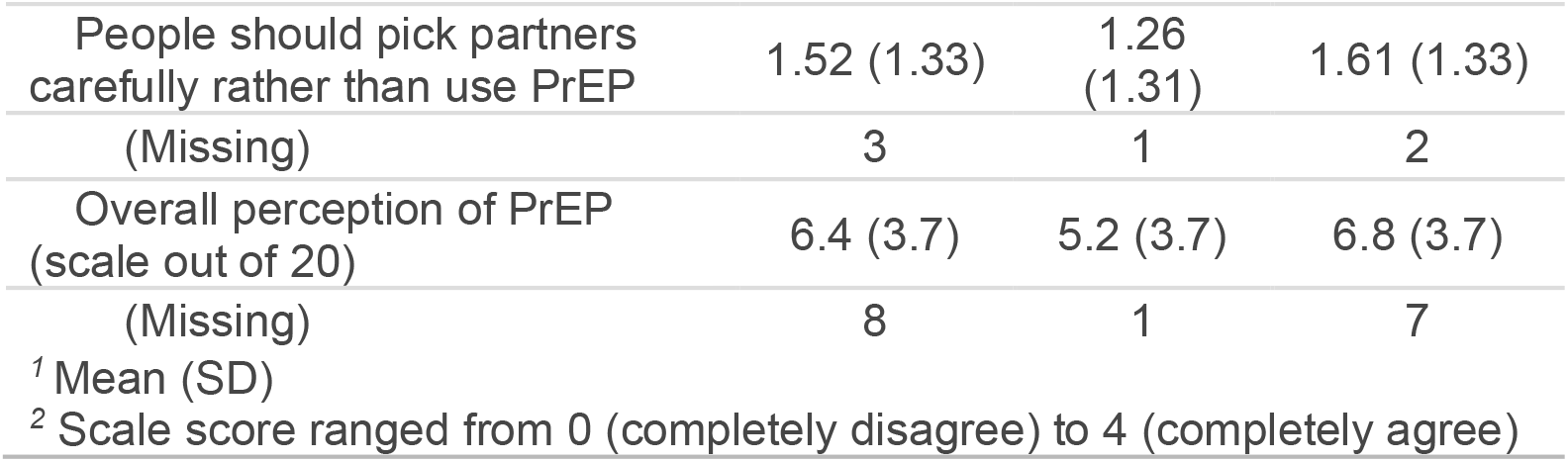
PrEP Stigmas and Perceptions.

### Characteristics Associated with Intention to Use PrEP

Overall, 118 (26%) reported they were highly likely/likely to use PrEP in the next year. In unadjusted analyses, there was a 35% increase in the odds of PrEP intention for each epidemiologic HIV risk factor (95% CI: [1.13, 1.62], p<0.01; Table 3). Participants with high HIV risk perception had 3.46 times the odds of PrEP intention compared to those with low HIV risk perception (95% CI: [2.07, 5.79], p<0.01). Participants who knew someone taking PrEP had 1.97 times the odds of PrEP intention compared to those who did not (95% CI: [1.21, 3.2], p<0.01). For each additional year of university, participants had a 52% increase in the odds of PrEP intention (95% CI: [1.26, 1.83], p<0.01). Those with prior PrEP use had 3.94 times the odds of PrEP intention (95% CI: [1.61, 9.6], p<0.01). Conversely, the odds of PrEP intention significantly decreased for every one unit increase in score on the negative PrEP perceptions scale (OR 0.88, 95% CI: [0.83, 0.94], p<0.010 and PrEP stigma scale (OR 0.91, 95% CI: [0.87, 0.95], p<0.01). Students’ financial status, whether the student had ever used condoms, their birth location, and whether they knew their partner’s HIV status were not associated with PrEP intention.

After adjusting for all factors that were significantly associated with PreP intention, except for perceived HIV risk (Table 3), participants with higher negative PrEP perception scores continued have reduced PrEP intention (aOR 0.9, 95% CI: [0.85, 0.97], p<0.01), as well as those with high negative PrEP stigma scores (aOR 0.92, 95% CI: [0.87, 0.97], p<0.01). The number of years a student had been in university continued to be associated with increased PrEP intention (aOR 1.48, 95% CI: [1.22, 1.8], p<0.01), though knowing someone else taking became marginally associated with PrEP intention (2.93, 95% CI: [0.97, 8.83], p = 0.06). Lastly, the number of epidemiologic HIV risk factors continued to be positively associated with PrEP intention (1.24, 95% CI: [1.01, 1.53], p = 0.04).

### Risk Perception as a Mediator in the Relationship Between Epidemiologic HIV Risk and PrEP Intention

The number of epidemiologic HIV risk factors was significantly associated with PrEP intention in both simple and adjusted logistic regression models (though without HIV risk perception included as a covariate; Table 3). However, when HIV risk perception was added to the adjusted model, the association between epidemiologic HIV risk and PrEP intention decreased in both magnitude and statistical significance, indicating that perceived HIV risk was a potential mediator between epidemiologic risk and PrEP intention (aOR: 1.24 dropping to 1.17, p-value: 0.04 increasing to 0.14, see Table 3). In this final model, the relationships between PrEP stigma, negative PrEP perceptions, year in university and PrEP intention were relatively unchanged from the previous adjusted model, though with the inclusion of self-perceived HIV risk, the association between having ever used PrEP and PrEP intention, having already been substantially weakened by the first set of covariates, basically disappeared (aOR 2.46, 95% CI: [0.83, 7.31], p = 0.11). Additionally, the relationship between perceived HIV risk and PrEP intention was the similar to the unadjusted model (aOR 3.08, 95% CI: [1.71, 5.55], p < .01). Lambda in Table 3 represents, for the final model, the proportion of the total variance of the aOR due to variance added by imputation of missing data: in this case, 28% of the variability in the estimate of the aOR of epidemiologic risk count on PrEP intention is due to multiple imputation. Analysis of the complete-case sample found similar associations except for a null association between the number of risk factors and PrEP intention (see Appendix).

Probabilistically speaking, a female student with one epidemiologic risk score, with the sample mean score for PrEP perception and PrEP stigma, who thinks her HIV risk is low, and has no other risk factors, has a hypothetical 13% probability of PrEP intention. If the same participant, however, believed she was at high risk of HIV, her probability of PrEP intention jumps to 32% (holding all other factors equal).

Given the change in the magnitude and significance of the association between epidemiologic risk count and PrEP intention after adjustment for perceived HIV risk, the association appears to be mediated by perceived HIV risk. Under a classic mediation framework, the odds ratio for the indirect effect of epidemiologic risk count on PrEP intention mediated by self-perceived HIV risk was 1.43 (Figure 1), and the proportion mediated of this relationship by self-perceived risk was 69%. These values were estimated with models that included PrEP stigma, PrEP perception, knowing a friend taking PrEP, and year in university as covariates.

**Figure 1.**
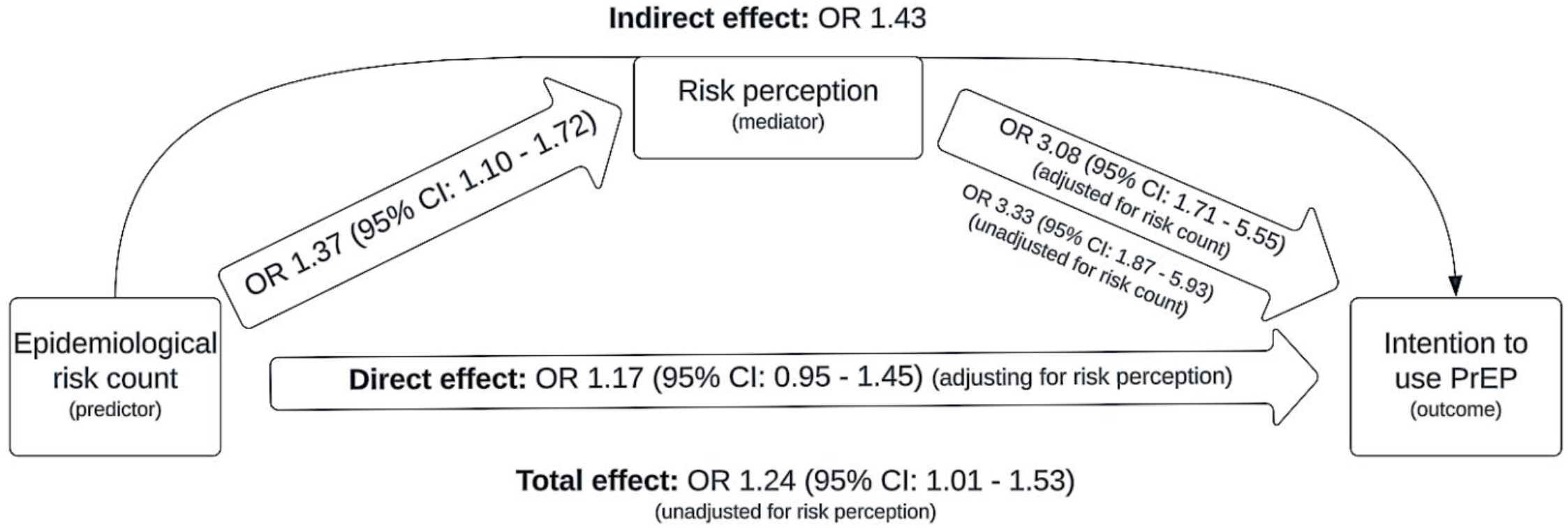
Results of Mediation Analysis (all modeling includes adjusted for relevant covariates)

### Alignment of Accurate Perceptions of HIV risk

Among those with low epidemiologic HIV risk, 86.1% accurately perceived their HIV risk to be low. However, among those with high epidemiologic HIV risk, only 28.6% accurately perceived their HIV risk to be high. McNemar’s test indicates participants do not rank themselves as high epidemiological risk at the same rate that they actually are high epidemiologic risk (maximum p-value from all tests < .001).

## DISCUSSION

This study contributes to the small body of research regarding female university students’ willingness to use PrEP in high HIV-prevalence African settings ^18,19^. Over one-quarter of female students in this study reported being highly likely/likely to use PrEP in the next year. The adjusted odds of PrEP intention were higher for those with less negative PrEP perceptions, less PrEP stigma, and perceived high risk for HIV. Thirty-eight percent of the sample met the criteria for being high HIV risk (i.e., ≥3 or more risk factors on the HIV Risk Index)^26^, but only 19% perceived they were high risk of acquiring HIV in the next year. A major contribution of this study is a better understanding of factors influencing intention to use PrEP among HIV at-risk female university students in Zambia, including the interconnected relationship of epidemiologic risk and perceived HIV risk.

Our findings indicate relatively high interest or PrEP intention but low actual use. We found that in our sample of sexually active female university students, 26% intended to use PrEP in the next year but less than 5% had or were currently using PrEP. A prior study from Zambia with both male and female university students who may or may not have been sexually active (n=346) found that 17% were willing to use PrEP and 4.9% had previously used PrEP ^19^; the sample did not stratify outcomes by gender. A study with male and female first year university students in Namibia found that 45% had heard of PrEP and, of those (n =104), 88% indicated PrEP intention with 8% prior PrEP use. ^46^ A study with male and female South African university students found 19% of participants were aware of PrEP, 15% knew where and how to get PrEP, and 2% had used PrEP. ^47^ In Lesotho, a study with female university students found that 32% were strongly willing to use PrEP if it were available in their community. ^18^ Taken together, this body of research, along with the present study findings, highlights the low use of PrEP among university students in several settings of Southern Africa, but opportunities to increase uptake given levels of interest.

In the present study, PrEP perceptions and PrEP stigma were significantly associated with participants’ intention to use PrEP in the next year. We found low levels of negative PrEP perceptions and PrEP stigma in this sample of young women, which is promising. However, some of the young women in this study continue to endorse PrEP stigma and negative PrEP perceptions, particularly that they would be judged if they used PrEP and PrEP causes risky sex. While still emerging, prior literature supports the finding that stigma is a barrier to PrEP use ^48-50^ Stigma may be especially problematic for PrEP use among AGYW whose identity is still forming as they transition to adulthood and have increased sensitivity to other’s opinions. ^50,51^

Our findings align with the conceptual framework by Hartmann et al. (2024) on PrEP stigma among AGYW in sub-Saharan Africa, including how it manifests, its intersection with other stigmas and vulnerabilities, and individual and health system outcomes. ^50^ Our study supports the recommendations of Hartmann et al. (2024) that advocate for PrEP stigma reduction and mitigation among AGYW through, for example, increasing community-wide knowledge dissemination of PrEP in non-stigmatizing ways, building resilience and social support, and offering counselling to address challenges with PrEP disclosure. ^50^ However, the authors do not specifically reference in-school AGYW or female university student populations. Our findings indicate that negative perceptions of PrEP and PrEP stigma are also barriers to PrEP intention among at-risk female university students. There are no effective interventions to our knowledge from Southern Africa that have successfully promoted PrEP uptake via addressing PrEP stigma/perceptions among AGYW.

In the present study, perceived high HIV risk emerged as a key factor associated with the odds of PrEP intention. Perceived HIV risk additionally was a significant mediator in the relationship between epidemiologic HIV risk and PrEP intention. A similar relationship was found by a Ugandan study that reported AGYW with higher “HIV Salience and Perception” scale scores were more likely to initiate and obtain PrEP refills through 6 months. ^52^ In Lesotho, a study with female university students reported increased willingness to use PrEP with perceived HIV risk.^18^ Similar to the present study, a study from Malawi reported that perceived HIV risk partially explained the relationship between epidemiologic HIV risk and PrEP interest among AGYW. ^27^ The prior research along with our findings highlight the importance of HIV risk perception and PrEP interest among AGYW, including female university students.

This study underscores the large, missed opportunity to prevent HIV among female university students at high risk of HIV acquisition who do not perceive themselves to be at risk. Similar to other studies from the region, we found that the female university students often underestimate their HIV risk. ^26,53^ In fact, over two-thirds (of the complete case subset) with high HIV risk in this study did not accurately perceive themselves to be at high HIV risk. There is an apparent need to align AGYW’s risk perception with their true HIV risk, including among female university students. Yet, the formation of HIV risk perceptions among AGYW is a complicated factor given that perceived risk is influenced by numerous internal (e.g., neurodevelopment, mental health) and external (duration, emotional attachment, and trust within a sexual relationship) factors, as well as biases including “optimism bias’’ where individuals tend to underestimate their own risk, and “present-bias” where individuals tend to focus on immediate rewards at the expense of long-term objectives. ^54,55^ Despite the complexity of this phenomena, there is an urgent need find effective ways to help AGYW better align their HIV risk perception with actual HIV risk. This will be critical for AGYW to make informed decisions not only about the use of oral PrEP, but also other PrEP products beginning to be available to African AGYW, such long-acting injections and vaginal rings. ^56^

### Limitations

The findings of this study should be interpreted within some limitations. The survey was based on a convenience sample of female university students who saw our advertisements and opted to take the online survey, making selection bias possible and not representative of all female university students in Zambia or other African higher education institutions. The study used self-report to collect data, which is vulnerable to social desirability and recall bias. This study was afflicted with a substantial amount of missing data, which was partly overcome with the aid of multiple imputation, still may have led to decreased accuracy for point estimates and wider confidence bands than what would be seen with complete data. Lastly, we recognize that PrEP intention does not always equate to PrEP uptake, and more studies are needed to examine PrEP uptake and persistence behaviors.

## Conclusion

This study found that intention to use PrEP among sexually active female university students in Zambia is affected by perceived HIV risk, along with PrEP perceptions and stigma. Perceived HIV risk mediates the relationship between epidemiologic HIV risk and PrEP intention. Among those with high epidemiologic HIV risk, most underestimated their risk. Along with education and stigma reduction, there is a need for effective approaches for female university students to accurately assess HIV risk and make informed decisions about PrEP use.

## Data Availability

All data produced in the present study are available upon reasonable request to the authors.

## APPENDIX 1

**Table 1:** Complete Case Regression Modeling Results.

| Variable | Simple models |  |  | Adjusted model |  |  | Final mediation model |  |  |
| --- | --- | --- | --- | --- | --- | --- | --- | --- | --- |
|  | OR | CI | p | aOR | CI | p | aOR | CI | p |
| # of risk factors | 1.20 | [0.92, 1.56] | 0.17 | 1.02 | [0.71, 1.45] | 0.93 | 0.98 | [0.65, 1.45] | 0.92 |
| PrEP stigma score | 0.91 | [0.86, 0.95] | < .01 | 0.97 | [0.87, 1.08] | 0.6 | 0.95 | [0.83, 1.07] | 0.39 |
| PrEP perception score | 0.88 | [0.83, 0.94] | < .01 | 0.82 | [0.7, 0.94] | < .01 | 0.88 | [0.75, 1.03] | 0.13 |
| Knows a friend taking PrEP | 2.05 | [1.25, 3.32] | < .01 | 2.29 | [0.63, 8.1] | 0.2 | 2.63 | [0.72, 9.25] | 0.13 |
| Poor financial situation | 0.60 | [0.31, 1.2] | 0.14 |  |  |  |  |  |  |
| Uses condoms | 1.12 | [0.69, 1.86] | 0.65 |  |  |  |  |  |  |
| Born in greater Zambia/international | 1.11 | [0.72, 1.7] | 0.65 |  |  |  |  |  |  |
| Year in university | 1.51 | [1.26, 1.83] | < .01 | 1.73 | [1.15, 2.76] | 0.01 | 1.57 | [1, 2.6] | 0.06 |
| Ever taken PrEP | 4.10 | [1.69, 10.31] | < .01 | 20.7 | [1.92, 319.91] | 0.02 | 9.34 | [0.95, 138.13] | 0.07 |
| Did not know partner's HIV status | 1.38 | [0.88, 2.15] | 0.15 |  |  |  |  |  |  |
| High self-perceived HIV risk | 4.20 | [2.49, 7.13] | < .01 |  |  |  | 4.01 | [0.98, 17.61] | 0.05 |

